# Ultrafast RNA extraction-free SARS-CoV-2 detection by direct RT-PCR using a rapid thermal cycling approach

**DOI:** 10.1101/2021.11.09.21265517

**Authors:** Robin Struijk, Anton van den Ouden, Brian McNally, Theun de Groot, Bert Mulder, Gert de Vos

## Abstract

The surging COVID19 pandemic has underlined the need for quick, sensitive, and high-throughput SARS-CoV-2 detection assays. Although many different methods to detect SARS-CoV-2 particles in clinical material have been developed, none of these assays are successful in combining all three of the above characteristics into a single, easy-to-use method that is suitable for large-scale use. Here we report the development of a direct RT-PCR SARS-CoV-2 detection method that can reliably detect minute quantities of SARS-CoV-2 gRNA in nasopharyngeal swab samples as well as the presence of human genomic DNA. An extraction-less validation protocol was carried out to determine performance characteristics of the assay in both synthetic SARS-CoV-2 RNA as well as clinical specimens. Feasibility of the assay and analytical sensitivity was first determined by testing a dilution series of synthetic SARS-CoV-2 RNA in two different solvents (water and AMIES VTM), revealing a high degree of linearity and robustness in fluorescence readouts. Following analytical performance using synthetic RNA, the limit of detection was determined at equal to or less than 1 SARS-CoV-2 copy/ul of sample in a commercially available sample panel that contains surrogate clinical samples with varying SARS-CoV-2 viral load. Lastly, we benchmarked our method against a reference qPCR method by testing 87 nasopharyngeal swab samples. The direct endpoint ultra-fast RT-PCR method exhibited a positive percent agreement score of 98.5% and a negative percent agreement score of 100% as compared to the reference method, while RT-PCR cycling was completed in 27 minutes/sample as opposed to 60 minutes/sample in the reference qPCR method. In summary, we describe a rapid direct RT-PCR method to detect SARS-CoV-2 material in clinical specimens which can be completed in significantly less time as compared to conventional RT-PCR methods, making it an attractive option for large-scale SARS-CoV-2 screening applications.

## Introduction

At the end of 2019, a novel strain of betacoronavirus called 2019_nCoV was identified^1^ in Wuhan, Hubei Province, China. In the following months, the virus proved highly contagious and as of September 20^th^, 2021, a total of 228,394,572 confirmed cases of COVID-19 including 4,690,186 deaths have been reported according to the World Health Organization Coronavirus (COVID-19) Dashboard. Shortly after its identification, 2019_nCoV was renamed to the current consensus^2^ term SARS-CoV-2 and the first complete SARS-CoV-2 RNA sequence was made public by the Chinese Centers for Disease Control and Prevention.

The emergence of SARS-CoV-2 has underlined the need for rapid and easy-to-implement diagnostic nucleic acid detection methods. Nucleic acid amplifications tests (NAAT) such as multiplex reverse-transcriptase polymerase chain reaction (RT-PCR) remain the test of choice^3^ for the detection of respiratory viruses due to their sensitivity, specificity, and time to virus detection. According to the CDC Interim Guidelines for Collecting and Handling of Clinical Specimens for COVID-19 Testing, nasopharyngeal swabs (NPS) are the preferred method to obtain biological samples for respiratory virus detection.

In this study we describe a highly efficient direct reverse transcriptase PCR (RT-PCR) protocol using our proprietary NextGenPCR ultra-fast thermocycler for the detection of SARS-CoV-2 genomic targets in NPS that does not require RNA extraction. For example to analyze 90 (?) samples from sample to result takes approximately 60 minutes with a total RT-PCR cycling time of 27 minutes. An input of only ≤ 10µl of patient material dissolved in viral transport medium (VTM) such as AMIES liquid is required to achieve a test result. Identification of SARS-CoV-2 positive and negative samples by end-point NextGenPCR RT-PCR is highly accurate as compared to a reference qPCR method on extracted RNA and, owing to its superior speed, is shown to be suitable for large-scale screening purposes where time to detection is a crucial factor.

## Methods

### Sample collection and preparation

#### Synthetic SARS-CoV-2 RNA

Logarithmic dilution series were made by serial dilution of SARS-CoV-2 RNA Control #2 (directed at the MN908947.3 viral RNA contingency [Wuhan-Hu-1 strain] Cat. no. #102024, Twist BioScience, USA) in nuclease-free H_2_O (cat. no. E476-500ML, VWR, Netherlands) or AMIES liquid (cat. no. K737B002VB, BioTRADING, Netherlands) to final concentrations as detailed in **Table 1**. Samples were kept on ice during workup to prevent RNA degradation as much as possible.

**Table 1.** Synthetic RNA dilution scheme.

| Sample name | Target concentration (copies/μl) | Dilution medium |
| --- | --- | --- |
| R1 H | $1 \times 10^4$ | Nuclease free H<sub>2</sub>O |
| R2 H | $1 \times 10^3$ | Nuclease free H<sub>2</sub>O |
| R3 H | $1 \times 10^2$ | Nuclease free H<sub>2</sub>O |
| R4 H | $1 \times 10^1$ | Nuclease free H<sub>2</sub>O |
| R5 H | $1 \times 10^0$ | Nuclease free H<sub>2</sub>O |
| R6 H | $1 \times 10^{-1}$ | Nuclease free H<sub>2</sub>O |
| R1 A | $1 \times 10^4$ | AMIES transfer medium |
| R2 A | $1 \times 10^3$ | AMIES transfer medium |
| R3 A | $1 \times 10^2$ | AMIES transfer medium |
| R4 A | $1 \times 10^1$ | AMIES transfer medium |
| R5 A | $1 \times 10^0$ | AMIES transfer medium |
| R6 A | $1 \times 10^{-1}$ | AMIES transfer medium |

### Surrogate clinical sample panel

Analytical performance of the SARS-CoV-2 and hRNaseP Primers and Probes, incl RT-PCR Chemistry 2× detection assay (cat. no. 50007, MBS, Netherlands) was determined using the SARS-CoV-2 Analytical Q Panel 01 (cat. no. SCV2AQP01-A, Qnostics Ltd, United Kingdom) described in **Table 2**. In addition to background human cells, the samples in this panel contain inactivated SARS-CoV-2 viral particles in transport medium of varying viral load. These samples thus closely mimic and are representative of human clinical samples as stated in manufacturer’s product description. Qnostics samples were thawed on ice and 4μl of each sample was directly pipetted in a 96×20μl MBS microplate in triplicate. Limit of detection (LOD) was determined by identification of the sample with the lowest concentration of viral copies that still produced a fluorescent signal higher than the negative control sample (SCV2AQP01-S09) in 3/3 replicates.

**Table 2.** Sample description and viral content in the SARS-CoV-2 Analytical Q Panel 01.

| Sample name | Target concentration (dC/ml) |
| --- | --- |
| SCV2AQP01-S01 | 1,000,000 |
| SCV2AQP01-S02 | 100,000 |
| SCV2AQP01-S03 | 10,000 |
| SCV2AQP01-S04 | 5,000 |
| SCV2AQP01-S05 | 1,000 |
| SCV2AQP01-S06 | 500 |
| SCV2AQP01-S07 | 100 |
| SCV2AQP01-S08 | 50 |
| SCV2AQP01-S09 | 0 |

### Nasopharyngeal swabs

A total of 101 individuals with suspected SARS-CoV-2 infection were sampled at the Canisius Wilhelmina Hospital (Nijmegen, The Netherlands) using Σ-Transwab® nasopharyngeal swabs (Copan Diagnostics, Italy). After collection, swabs were directly transferred to a sterile microtube containing 1mL AMIES liquid and heat-inactivated at 100°C for 10 minutes. An aliquot was taken from the tube containing the swab and subjected to RNA extraction followed by qPCR measurements (reference method), and the remaining volume was shipped to our laboratory in Goes where it was subjected to direct RT-PCR as described below approximately 24-48h after primary sampling.

### Polymerase chain reactions

#### NextGenPCR direct RT-PCR method

Our SARS-CoV-2 direct RT-PCR method using the NextGenPCR thermal cycler (MBS, The Netherlands) is based on multiplex RT-PCR amplification of three target sequences followed by end-point detection of amplicon-specific probe fluorescence to determine the presence of SARS-CoV-2 gRNA and human gDNA material in a sample. In the reaction, FAM-labelled (fluorescein amidites) oligonucleotide probes are used to detect SARS-CoV-2 N1 and ORF1ab gene sequences and Cy5-labeled oligonucleotide probes are used to detect a sequence in the human RPP30 gene. Primers/probes specific for RPP30 were included as an internal control target, serving both to confirm absence of PCR inhibition and indicate the presence of human genomic material in sample wells, aiding laboratory personnel to verify that sampling and pipetting went correctly. Primer and probe oligonucleotide sequences were designed using extensive bioinformatical analyses and in silico digital PCR to minimize amplification of off-target sequences and formation of primer-dimers or other secondary structures. Cross-reactivity of the reagents with other respiratory viruses was evaluated and showed no amplification of targets in off-target organism genomic sequences (data not shown). A total of 16μl of RT-PCR master mix containing 10μl RT-PCR Chemistry 2x, 1.6μl SARS-CoV-2/hRNaseP Primers and Probes and 4.4μl nuclease-free H_2_O (MBS, Netherlands) was added to 4μl of sample resulting in a total reaction volume of 20µl. The microplate was heat-sealed after pipetting with a transparent Clear Heat Seal (MBS, Netherlands) on a NextGenPCR Semiautomatic Heat Sealer (MBS, Netherlands) and transferred to a NextGenPCR machine for thermal cycling using the RT-PCR program detailed in **Table 3**. Three wells containing Human Positive Control material from the MBS detection assay were included to confirm efficient PCR cycling. After PCR was completed, the sealed microplate was snapped to an imaging anvil (MBS, Netherlands), transferred to a FLUOstar Omega Microplate Reader (BMG Labtech, Germany), scanned and fluorescence readout results exported to Excel for downstream data analysis. For measurements in synthetic RNA, the microplate was also scanned on a Bio-1000F Gel Imager (Microtek, Taiwan) and the results interpreted using custom designed QuickDetect software (MBS, Netherlands).

**Table 3.** NextGenPCR reverse transcriptase RT-PCR and qPCR cycling programs.

|  | NextGenPCR |  |  | qPCR (LC480-II) |  |  |
| --- | --- | --- | --- | --- | --- | --- |
| Program step | Cycle no. | Temp. (°C) | Time (s) | Cycle no. | Temp. (°C) | Time (s) |
| Reverse transcription | 1 | 55 | 300 | 1 | 50 | 300 |
|  |  | 98 | 60 |  | 95 | 20 |
| Initial amplification | 5 | 98 | 10 | No initial amplification |  |  |
|  |  | 60 | 20 |  |  |  |
|  |  | 72 | 3 |  |  |  |
| Amplification | 45 | 98 | 5 | 45 | 95 | 3 |
|  |  | 60 | 12 |  | 60 | 30 |
|  |  | 72 | 3 |  |  |  |
| Total time (s) | 1425 |  |  | 3597 |  |  |

### Reference qPCR method

The SARS-CoV-2 qPCR detection method described by Corman and colleagues^4^ was used as the reference method for this study. In short, nucleic acids were extracted from the sample/AMIES mixture using a MagNA Pure 96 DNA And Viral NA Small Volume Kit on a MagNA Pure 96 Instrument (both Roche Diagnostics GmbH, Mannheim, Germany) according to manufacturer’s instructions. Using Xiril robotic workstations (Roche Diagnostics GmbH, Mannheim, Germany), an internal control sequence specific for phocine distemper virus (PhDV) was added to the sample prior to nucleic acid extraction. After extraction, 5 μl TaqMan Fast Virus 1-Step Master Mix (ThermoFisher Scientific) and 5 μl primers and probes (**Table 4**) were added to 10 μl of spiked-in sample. Thermal cycling was performed on a LC480-II instrument (Roche) using the RT-PCR program detailed in **Table 3**. Data analysis was performed using FLOW software (Roche) and a threshold Ct-value of 40 was used as cut-off to interpret results as positive (Ct ≤ 40), negative (no Ct after 50 cycles) or indeterminate (Ct between 40-50).

**Table 4.**
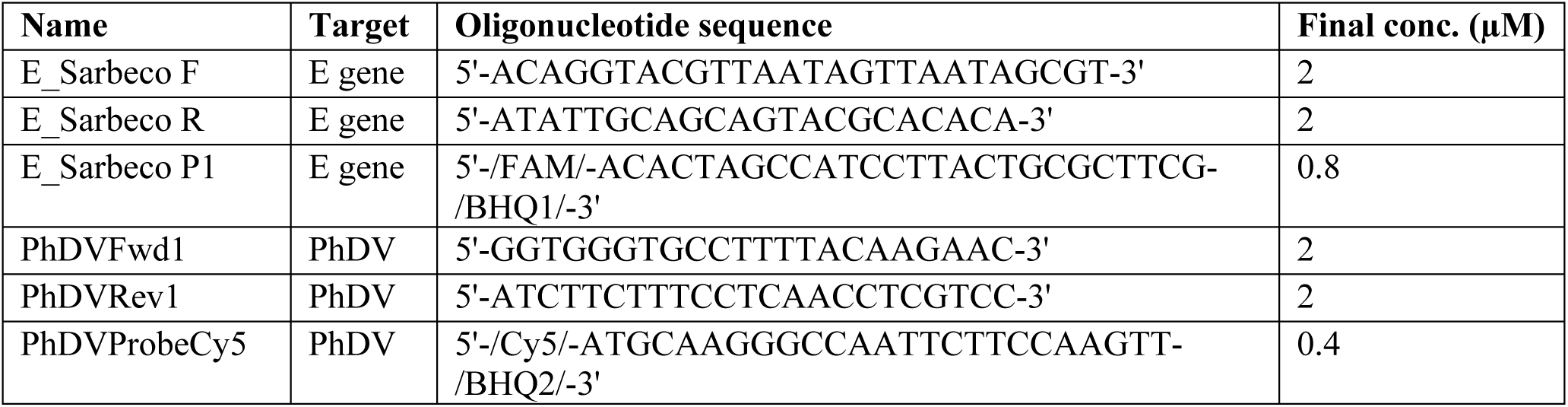
Primers used in the reference qPCR SARS-CoV-2 detection method.

## Results

### Direct RT-PCR on synthetic RNA dissolved in water or AMIES liquid shows similar amplification linearity and limit of detection

We first determined the analytical sensitivity of our assay in a serial logarithmic dilution series of synthetic RNA (**Table 1**). Samples were diluted in nuclease-free water to establish a baseline reading, as well as AMIES liquid to detect possible discrepancies of the fluorescence readouts using an actual VTM. A sample was called as being detected when all three replicates showed a significantly higher fluorescence level (sample RFU ≥ mean RFU in NTC + 3 × standard deviation) as compared to background signal in non-template control (NTC) wells within the same medium.

Using the above cut-offs, FAM fluorescence was detected on a Bio-1000F blue light scanner in synthetic RNA dilutions ranging from 1 to 10,000 SARS-CoV-2 copies/µl in both solvents, but not in the 0.1 copies/µl sample (**Figure 1A**). Dilutions made in nuclease-free H_2_O as compared to AMIES liquid were shown to have slightly higher R^2^-values (R^2^ = 0.91 and R^2^ = 0.87 in nuclease-free water and AMIES liquid, respectively) as well as a higher signal amplitude when measured on a FLUOstar Omega microplate reader, most noticeable in samples with a SARS-CoV-2 concentration ≥ 100 copies/µl (**Figure 1B**). The FLUOstar Omega is equipped with multiple single channel fluorescent detectors allowing for qualification of FAM signals as well as CY5 signals, the latter of which cannot be detected with a Bio-1000F blue light scanner. Human *RPP30* was detected in the kit positive control only (p < 0.05, two-sided Student’s t-test), indicating that the used primer oligonucleotides are specific for the intended *RPP30* human genomic target without off-target amplification in samples lacking *RPP30* templates (**Figure 1C**).

**Figure 1.**
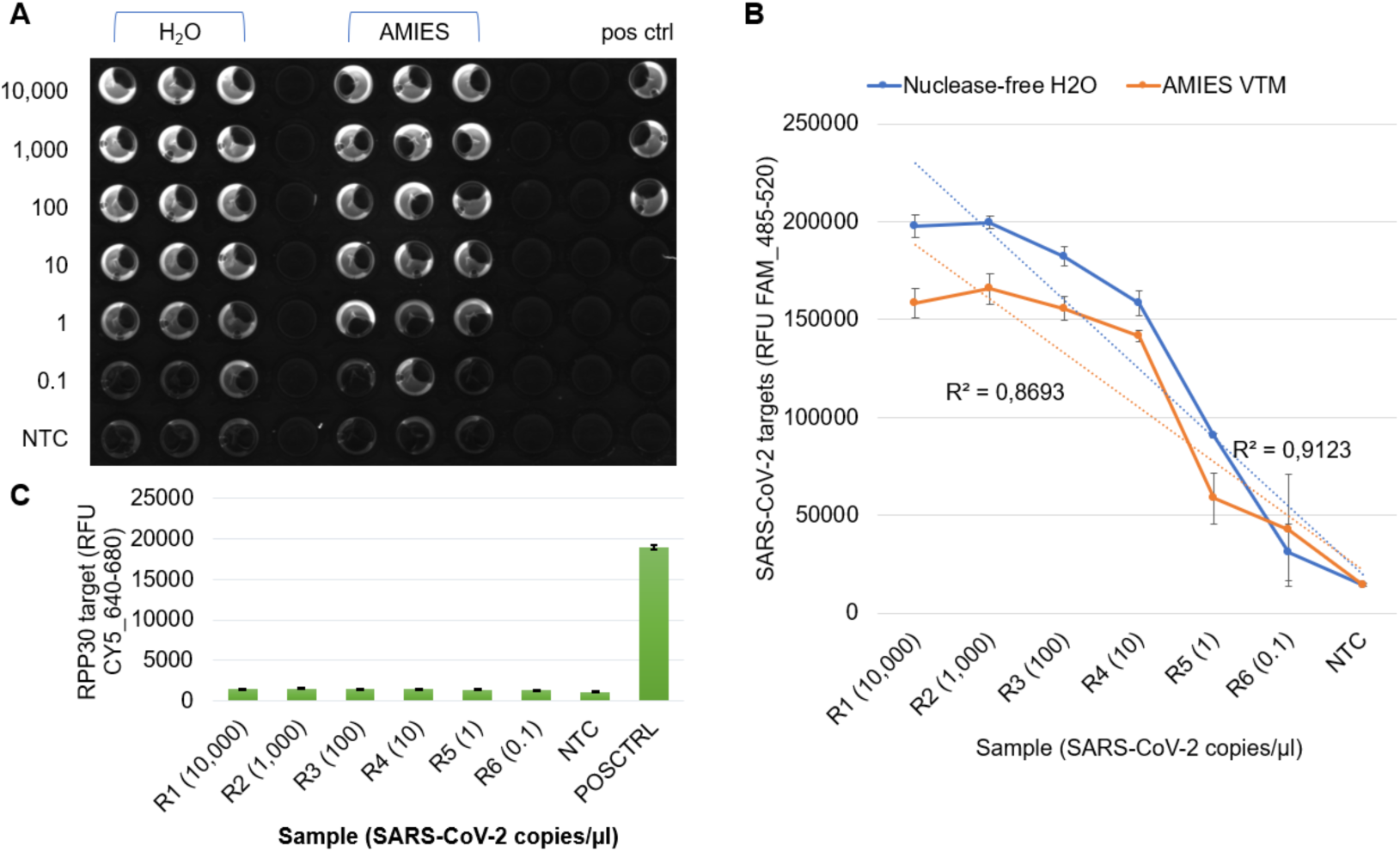
Analytical sensitivity of the NextGenPCR SARS-CoV-2 detection assay as determined in a serial logarithmic dilution series of synthetic SARS-CoV-2 RNA. (A) Image of the microplate generated by a blue-light imager show clear distinction between samples with FAM fluorescence and samples without fluorescence. (B) Relative fluorescence units (RFU) measured on the FLUOstar Omega plate reader follows a linear profile with decreasing concentrations of synthetic RNA. Whiskers represent standard error of mean. (C) RPP30 is only detected in the positive control and not in synthetic RNA samples.

### Direct RT-PCR on a panel of surrogate clinical samples provides accurate and sensitive SARS-CoV-2 detection without prior nucleic acid extraction

Next, a commercially available analytical sample panel containing samples with a known concentration of inactivated SARS-Cov-2 particles (ranging from 1 × 10^6^ to 5 × 10^1^ copies per milliliter of sample) mixed with a human cell matrix was tested using the direct RT-PCR protocol (**Table 2**). SARS-CoV-2 targets were detected in samples SCV2AQP01-S01 through SCV2AQP01-S05 indicating a limit of detection of ≤ 1 copies/µl sample (**Figure 2A**). One out of three replicates in sample SCV2AQP01-S06 were detected and SARS-CoV-2 targets were not detected in samples SCV2AQP01-S07 through SCV2AQP01-S09. The internal *RPP30* control target was detected in every well, confirming presence of human genomic DNA in all samples (**Figure 2B**). The data showed a linear decrease in abundance of fluorescence of SARS-CoV-2 targets with decreasing viral load in the sample set (R^2^ = 0.89, data not shown).

**Figure 2.**
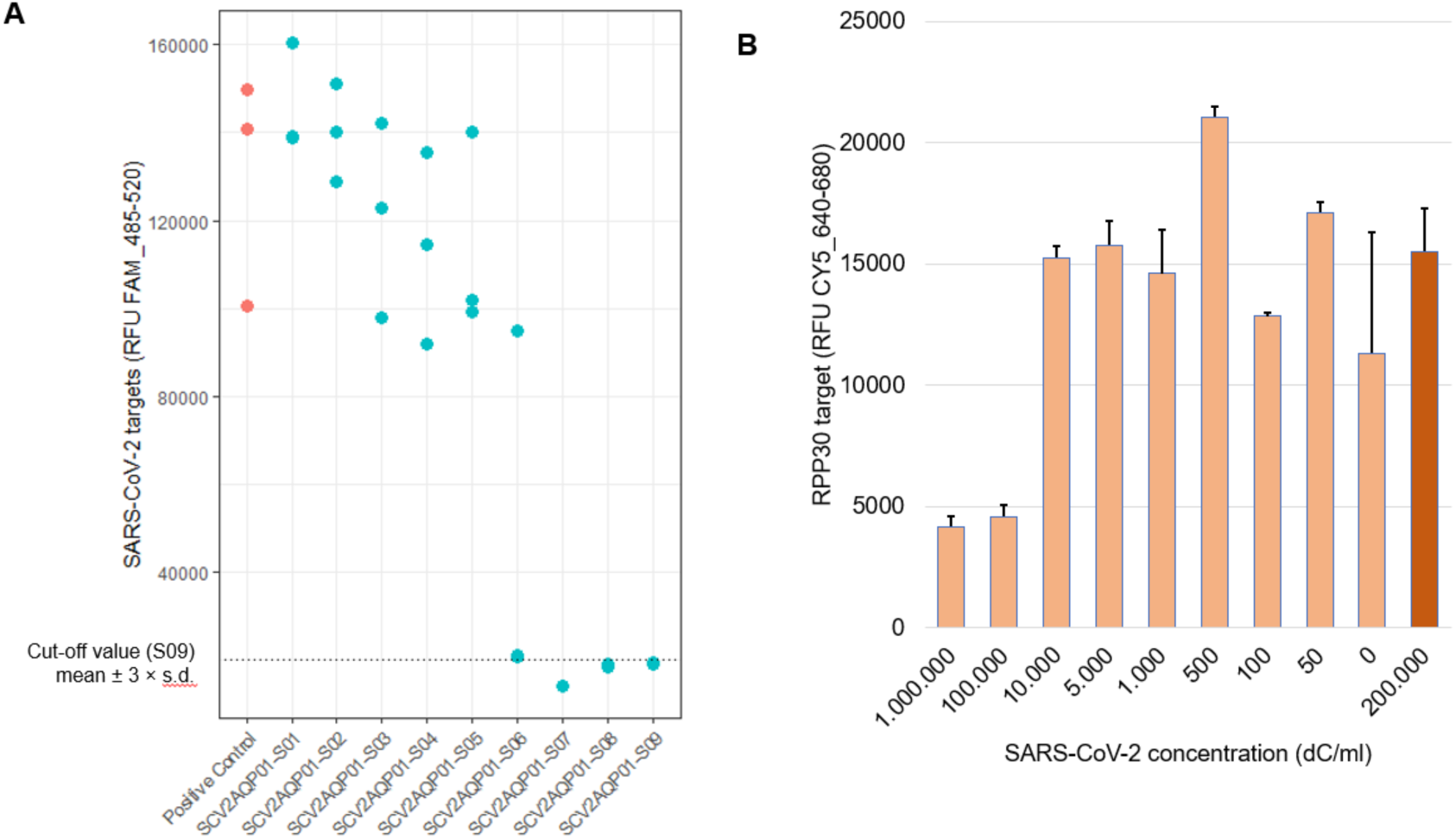
Fluorescence values for samples SCV2AQP01-S01 through SCV2AQP01-S09 of the Qnostics validation sample panel measured in triplicate. (A) ORF1ab and N1 (SARS-CoV-2 targets) are detected through FAM-labeled probes and (B) human RPP30 (internal control) is detected through a CY5-labeled probe. Bars represent mean relative fluorescence units (RFU), whiskers indicate standard error of mean.

### Direct RT-PCR demonstrates high clinical sensitivity, specificity and sample calling agreement as compared to reference qPCR methodology

Finally, clinical sensitivity and specificity were determined using direct RT-PCR on 87 nasopharyngeal swab samples that were identified as positive or negative for SARS-CoV-2 with a reference qPCR method targeting the *E* gene that is recommended to use as a first-line screening tool in the original article describing the method^4^. Of these samples, 64 were tested positive and 23 were tested negative for SARS-CoV-2. As additional controls, we included 17 non-template controls (NTC) with only AMIES viral transport medium as input, 6 non-template controls with only H_2_O as input and 3 positive controls containing plasmid SARS-CoV-2 *N1* sequences only.

SARS-CoV-2 target sequences were detected in 64/64 positive samples and 1/23 negative samples, while *RPP30* sequences were detected in 85/87 samples corresponding to a false negative detection rate of 2.3% for *RPP30* detection in clinical samples (**Figure 4A**). Both samples with undetectable *RPP30* were positive for SARS-CoV-2 and were likely undetected due to competition for PCR reagents between primer sets, a phenomenon that is well described in multiplex PCR which we could also observe here (see **Figure 2**, reduced CY5 label fluorescence in samples with high SARS-CoV-2 copy number). We observed strong correlation between fluorescence readouts following direct RT-PCR and threshold cycle (C_t_) numbers with a correlation coefficient of R^2^ = 0.84 and concordance statistic of ρ_c_ = 0.79 between the two techniques (**Figure 3**). A clinical agreement study was carried out to estimate percent positive agreement (PPA, or clinical sensitivity) and percent negative agreement (NPA, or clinical specificity) scores as recommended by the FDA in the “CLSI EP12: User Protocol for Evaluation of Qualitative Test Performance” protocol. As detailed in **Figure 4B**, PPA was calculated at 98.5% and NPA was calculated at 100%.

**Figure 3.**
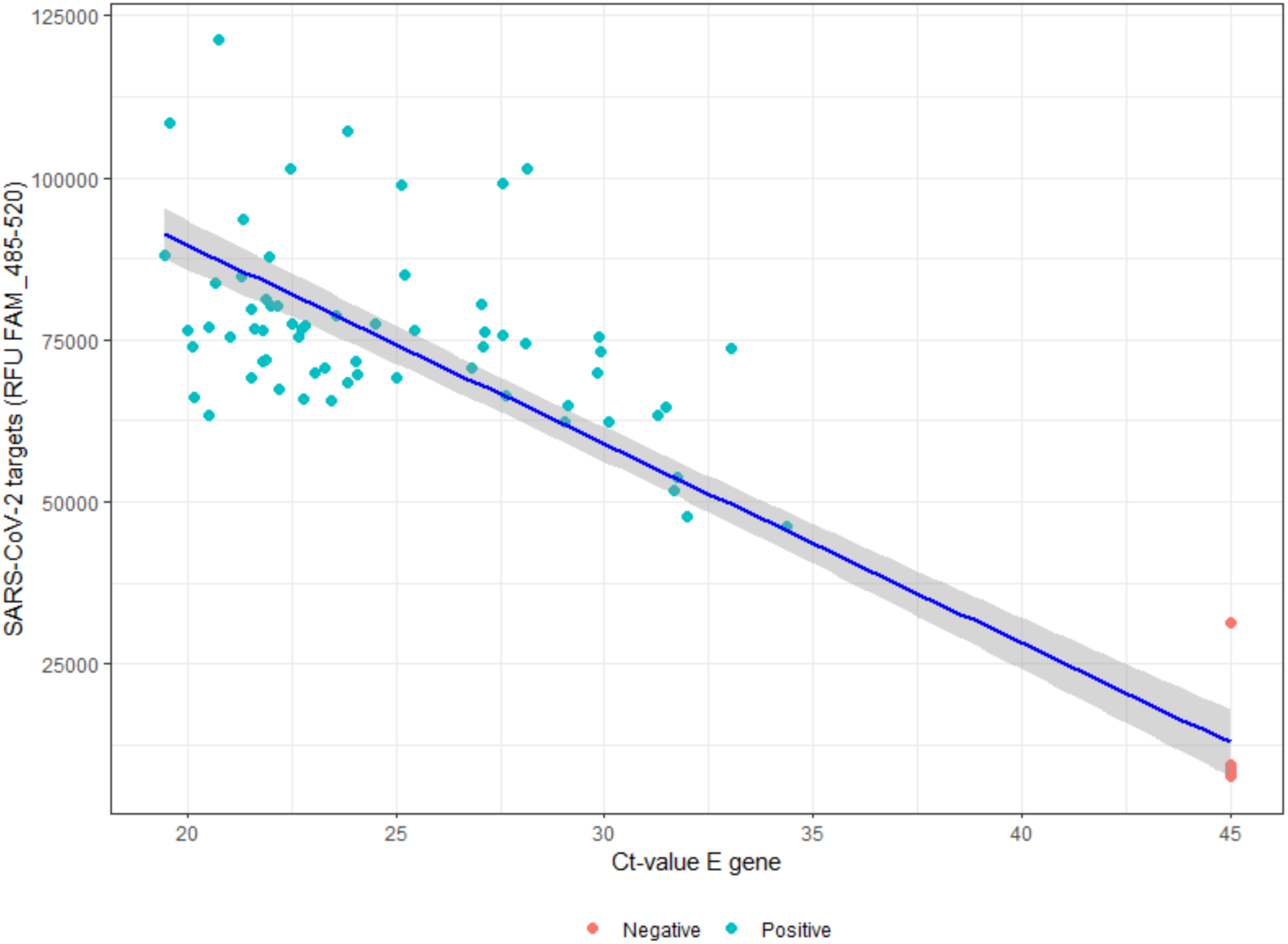
Comparison of the experimental NextGenPCR detection method versus an established qPCR reference method. Fluorescence levels detected by the experimental method show a linear correlation with Ct-values and show a distinct difference between positive and negative samples.

**Figure 4.**
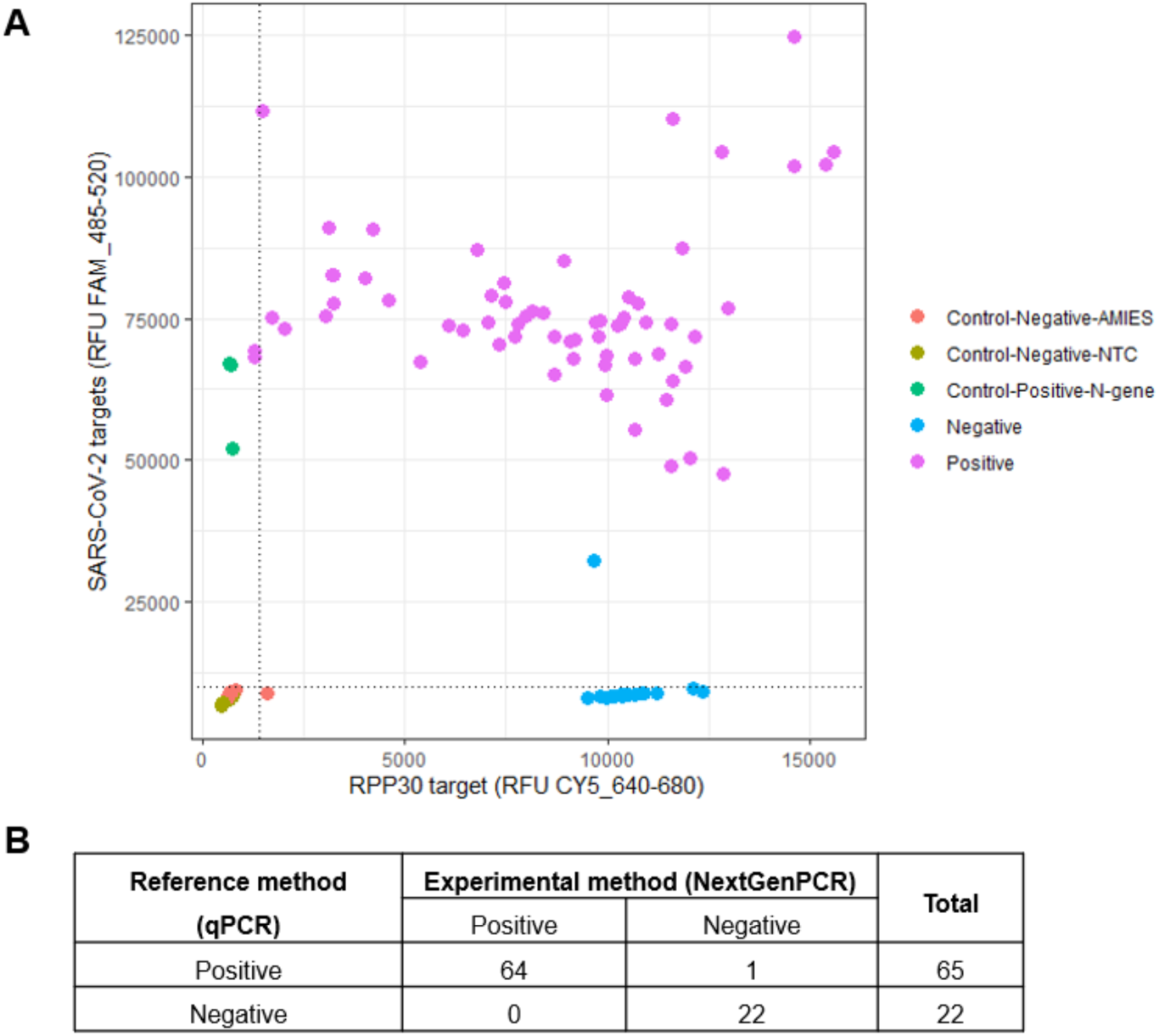
Distribution of fluorescence signals of the SARS-CoV-2 targets (y-axis) and RPP30 target (x-axis) for all included samples. Cut-off values are provided for positive SARS-CoV-2 signals (horizontal black line, RFU [640-680] = 1436) and positive RPP30 signals (vertical black line, RFU [485-520] = 9985).

### Direct RT-PCR on an additional 14 clinical samples positive for respiratory viruses other than SARS-CoV-2 confirms NextGenPCR primer-probe specificity

To confirm analytical specificity of the used primer and probes in the NextGenPCR direct RT-PCR method, 14 additional clinical specimens with varying respiratory viral infections other than SARS-CoV-2 (see **Table 5**) were analyzed on a BioRad CFX96 qPCR system in addition to NextGenPCR analysis as described above. No off-target amplification of FAM-labeled fragments was observed in these samples (CT undetermined on CFX96, NextGenPCR RFU ranging from ∼16,000 to 20,000 in the FAM channel with a signal of 18,000 RFU in the negative SPEC0004 sample), except for three samples where 400 copies/µl of MBS SARS-CoV-2/RPP30 positive control material was spiked-in prior to sample workup to control for potential PCR inhibition due to buffer components (**Figure 5**). The signal amplitude in the spike-in samples averaged to 142,707 RFU in the FAM channel, well above the average FAM signal (RFU 76,667) in the 64 positive samples described above.

**Table 5.** NextGenPCR assay specificity analysis of 14 patient samples with different respiratory virus infections other than SARS-CoV-2. RSV indicates respiratory syncytial virus; InfA, influenza A; InfB, influenza B; HMPV, human metapneumovirus; EV, enterovirus; AV, adenovirus; SARS-CoV-2, severe acute respiratory syndrome coronavirus 2. MBS positive control (400 copies/µl) was used as spike-in (-sp) for samples 1, 11 and 13.

| Sample | Pathogen | LC480-II | CFX96 |  | NextGenPCR |  |
| --- | --- | --- | --- | --- | --- | --- |
|  |  | C<sub>T</sub>-pathogen | C<sub>T</sub>-FAM | C<sub>T</sub>-CY5 | RFU-FAM | RFU-CY5 |
| SPEC0001 | RSV | 29.93 | NA | 26.12 | 16834 | 19084 |
| SPEC0002 | InfB | 35.65 | NA | 23.33 | 17958 | 19733 |
| SPEC0003 | RSV | 23.95 | NA | 26.7 | 17010 | 19502 |
| SPEC0004 | None | NA | NA | 21.33 | 18006 | 20542 |
| SPEC0005 | RSV | 38.18 | NA | 24.67 | 17446 | 18850 |
| SPEC0006 | InfB | 35.75 | NA | 26.09 | 18943 | 21102 |
| SPEC0007 | InfB | 33.16 | NA | 26.89 | 18821 | 20435 |
| SPEC0008 | InfA | 25.32 | NA | 20.54 | 19431 | 20510 |
| SPEC0009 | InfA | 24.3 | NA | 25.04 | 17676 | 20674 |
| SPEC0010 | InfA | 23.96 | NA | 22.2 | 17947 | 21715 |
| SPEC0011 | HMPV | 31.18 | NA | 23 | 17506 | 19973 |
| SPEC0012 | HMPV | 34.38 | NA | 21.84 | 17862 | 20455 |
| SPEC0013 | EV | 21 | NA | 30.93 | 17498 | 20288 |
| SPEC0014 | AV | 35 | NA | 21.88 | 20190 | 20559 |
| SPEC0001-sp | RSV + SARS-CoV-2 | NA | 30.31 | 27.23 | 145080 | 18762 |
| SPEC0011-sp | HMPV + SARS-CoV-2 | NA | 29.67 | 29.58 | 157763 | 19984 |
| SPEC0013-sp | EV + SARS-CoV-2 | NA | 29.91 | 24.35 | 133763 | 18628 |
| MBS_POS | SARS-CoV-2 | NA | 29.19 | 28.87 | 142544 | 17220 |
| MBS_POS | SARS-CoV-2 | NA | 29.01 | 29.25 | 147992 | 18038 |
| MBS_POS | SARS-CoV-2 | NA | 28.32 | 28.62 | 137585 | 15775 |
| NTC | None | NA | NA | NA | 13740 | 1044 |
| NTC | None | NA | NA | NA | 13884 | 986 |
| NTC | None | NA | NA | NA | 13593 | 977 |

**Figure 5.**
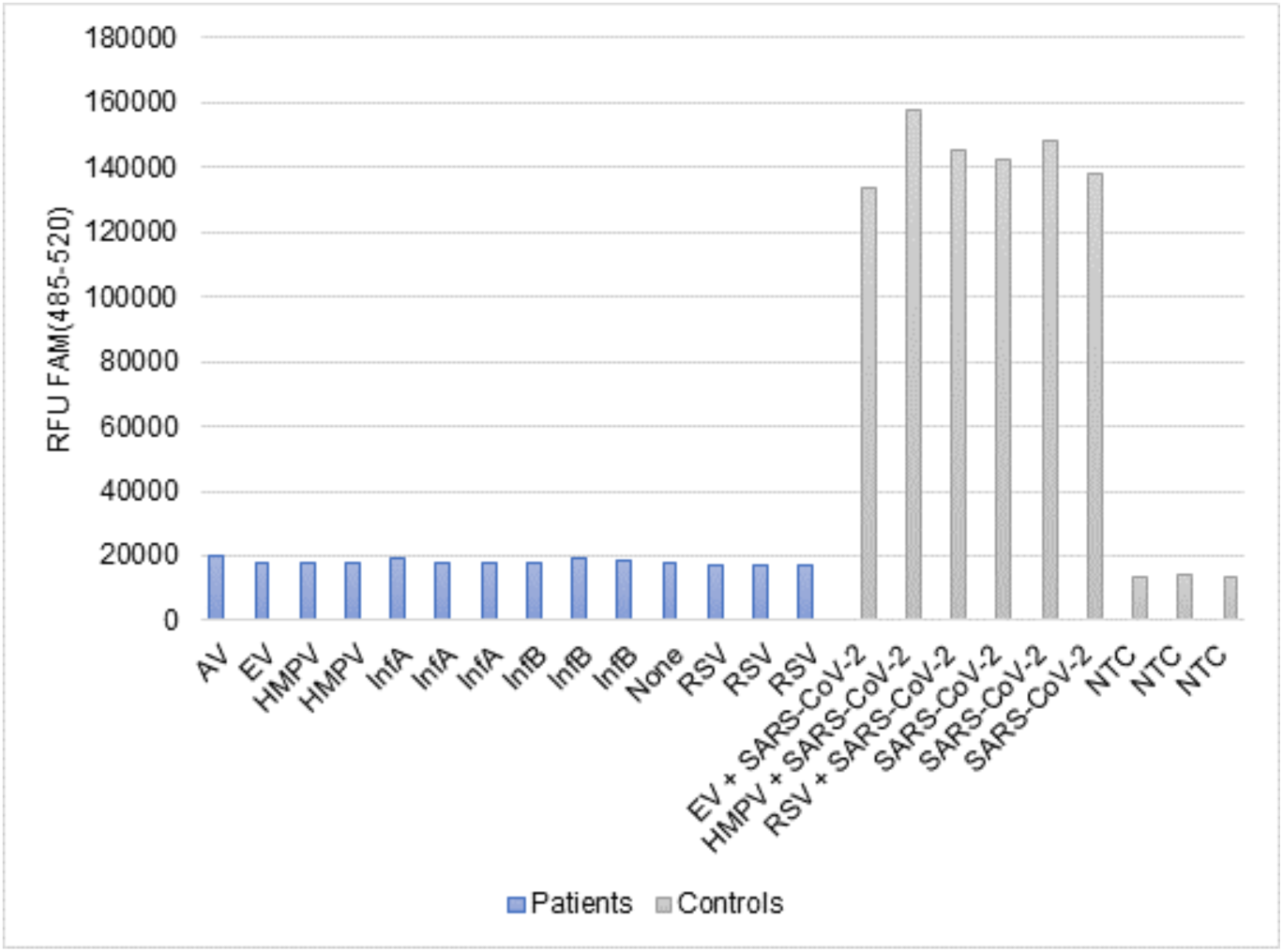
Relative fluorescence units (RFU) measured in the FAM channel for SARS-CoV-2 targets in clinical samples with a respiratory viral infection other than SARS-CoV-2. Blue bars represent clinical specimens, gray bars represent control samples. Samples SPEC0001, SPEC0011 and SPEC0013 were spiked (-sp) with MBS positive control plasmids to a final concentration of 400 copies/µl.

## Discussion

The development of screening and diagnostic tools to detect SARS-CoV-2 sequences in clinical specimens is of global importance. To reduce overall runtime and increase sample throughput, a substantial number of publications describe the use direct RT-PCR testing in primary clinical samples without prior nucleic acid extraction^5, 6^. In this paper, we describe the use of the NextGenPCR SARS-CoV-2 detection chemistry and NextGenPCR thermal cycler to accurately detect SARS-CoV-2 RNA in clinical samples with significantly reduced PCR cycling time and without the need for sample lysis or nucleic acid extraction. The analytical sensitivity of our developed assay was determined at 1.0 × 10^0^ copies/ul. The same limit of detection was observed using direct RT-PCR in the Qnostics sample panel, confirming that clinical samples with a SARS-CoV-2 viral load of ≥ 1 copy/ul can be consistently detected in our assay. When compared to a reference quantitative RT-PCR method targeting the SARS-CoV-2 E gene, our assay accurately identified samples in which SARS-CoV-2 RNA sequences are present as evidenced by the high clinical agreement scores between the two techniques (PPA = 98.5% and NPA = 100%).

The choice for a multiplex dual probe design targeting the SARS-CoV-2 *ORF1AB* and *N1* targets with the same fluorophore (FAM) and the human *RPP30* target with a second fluorophore (CY5) was made based on several theoretical advantages. The main contribution of a dual amplicon detection setup is an increased signal amplitude generated for the SARS-CoV-2 targets, since both amplicons contribute to the fluorescent signal generated in the sample instead of one. Additionally, a dual probe design provides strong analytical protection against sequence variation of the target sequences, which is relevant due to the significant rate at which SARS-CoV-2 variants are detected in the general population^7^. Currently, the capacity to detect SARS-CoV-2 has not been impacted by recently identified variants such as B.1.1.7 (British) and B.1.351 (South Africa) since these variants contain mutations^8^ in the S gene and our assay targets different sequences of the SARS-CoV-2 genome. It is essential that we and other assay developers stay vigilant in variant surveillance due to fast and frequents mutation of SARS-CoV-2 and ensure that SARS-CoV-2 sequence detection capacity does not deteriorate as time progresses due to mutational drift. To optimize the efficacy of our assay, we opted for thermal lysis of NPS specimens at 98°C prior to RT-PCR based on previous reports where improved detection rates of *N1* target sequences have been reported^5^ in a singleplex direct RT-PCR application. The beneficial effect of thermal lysis was confirmed by a second study^9^ that compared five RT-PCR master mixes for SARS-CoV-2 detection by multiplex RT-PCR describing improved detection rates of *N1* and *N2* target sequences after thermal sample lysis at 98°C (81%) as compared to lysis at 65°C (56%) or no thermal lysis (52%) prior to RT-PCR testing.

Apart from SARS-CoV-2 testing, the NextGenPCR thermal cycling system is also suitable for the detection of other pathogens including other microorganisms (viruses, bacteria) and pathogenic genetic variants in the human genome. Brons and colleagues^10^ describe the use of NextGenPCR for fast detection of multiple uropathogenic *Escherichia coli* strains in urinary tract infection patients in 52 minutes, significantly lowering the turnaround time over currently used techniques.

## Concluding remarks

In summary, we here present a screening method for the detection of SARS-CoV-2 viral particles in human clinical material through direct RT-PCR. Owing to ultrafast thermal cycling capacity, the NextGenPCR system may aid to reduce the typical turnaround time of sample to result from ∼4-6 hours in conventional RT-PCR to ∼45 minutes with NextGenPCR (RT-PCR cycling time of 27 minutes). With lower TAT an overall increased laboratory throughput of sample tests can be achieved characterized by a high level of analytical accuracy. This makes NextGenPCR highly suitable for large-scale screening applications that may benefit public spaces where many individuals are tested for SARS-CoV-2.

## Data Availability

All data produced in the present study are available upon reasonable request to the authors

## Acknowledgements

We would like to thank Martin Donker (Isogen Life Sciences B.V., Netherlands) for kindly providing an aliquot of the SARS-CoV-2 Analytical Q Panel 01.

